# Rapid Identification of Bacterial isolates Using Microfluidic Adaptive Channels and Multiplexed Fluorescence Microscopy

**DOI:** 10.1101/2023.07.16.23292615

**Authors:** Stelios Chatzimichail, Piers Turner, Conor Feehily, Alison Farrar, Derrick Crook, Monique Andersson, Sarah Oakley, Lucinda Barrett, Hafez El Sayyed, Jingwen Kyropoulos, Christoffer Nellåker, Nicole Stoesser, Achillefs N. Kapanidis

**Affiliations:** Department of Physics, University of Oxford, Parks Road, Oxford, OX1 3PJ, United Kingdom; Kavli Institute for Nanoscience Discovery, University of Oxford, South Parks Road, Oxford OX1 3QU, United Kingdom; Nuffield Department of Medicine, University of Oxford, John Radcliffe Hospital, Oxford, OX3 9DU, United Kingdom; Department of Microbiology and Infectious Diseases, Oxford University Hospitals NHS Foundation Trust, Oxford, OX3 9DU, United Kingdom; Nuffield Department of Women’s & Reproductive Health, University of Oxford, Big Data Institute, Oxford, OX3 7LF, United Kingdom

## Abstract

We demonstrate the rapid capture, enrichment, and identification of bacterial pathogens using Adaptive Channel Bacterial Capture (ACBC) devices. Using controlled tuning of device backpressure in polydimethylsiloxane (PDMS) devices, we enable the controlled formation of capture regions capable of trapping bacteria from low cell density samples with near 100% capture efficiency. The technical demands to prepare such devices are much lower compared to conventional methods for bacterial trapping and can be achieved with simple benchtop fabrication methods. We demonstrate the capture and identification of seven species of bacteria with bacterial concentrations lower than 1000 cells/mL, including common Gram-negative and Gram-positive pathogens such as *Escherichia coli* and *Staphylococcus aureus*. We further demonstrate that species identification of the trapped bacteria can be undertaken in the order of one-hour using multiplexed 16S rRNA-FISH with identification accuracies of 73-99% with unsupervised classification methods.

## Introduction

Antibiotics are critical to the treatment of bacterial infections, but their widespread use has contributed to the emergence of bacterial strains that can survive antibiotic treatment. These antibiotic-resistant bacteria pose a serious public health threat, causing more than 1.27 million deaths per year worldwide,^1^ and threaten a return to the pre-antibiotic era.

One of the means of controlling antibiotic resistance is by improving the accuracy and turnaround of diagnostic tests that detect the presence of bacteria and define their susceptibility to bacterial treatment directly from clinical samples. Culture-based tests for bacterial identification and antimicrobial resistance (AMR) already exist, but typically require 1-2 days to complete from the time the specimen is obtained from the patient. During that time, and especially for severe cases of infection, healthcare professionals prescribe broad-spectrum antibiotics based mainly on epidemiological data, when these are available. To address this, there is a clear need for rapid diagnostic assays that can be applied directly to clinical samples (i.e., without the need to first prepare clinical isolates, i.e., pure, monomicrobial cultures) to determine the type of infecting bacteria and enable accurate and rapid antibiotic susceptibility testing to guide patient treatment and antimicrobial stewardship.

A promising approach for rapid bacterial identification and analysis from clinical samples involves microscopy techniques. While detection of bacterial pathogens using microscopy on clinical isolates is relatively simple, detection in complex clinical samples is much more challenging, since, in addition to the need to distinguish bacteria from the sample matrix, some clinical samples have low bacterial concentrations in the absence of pre-culture steps (e.g., 1-10 colony-forming units (CFUs)/mL for blood).^2,3^ To enable the study and quantification of bacteria by microscopy, it is thus important that the target bacteria are enriched and spatially concentrated so as to allow rapid identification and interrogation of the pathogen.^4^

To achieve bacterial enrichment, researchers have previously used a combination of size-selection modules such as nano-porous monoliths,^4^ hydrodynamic traps,^5^ and nanostructured channels,^6^ as well as bacterial surface affinity agents, such as Apolipoprotein-H and Mannose-binding lectin.^7,8^ Further distinguishing features, such as small differences in bacterial length and morphology, have also been leveraged in combination with viscoelastic focusing to enrich bacterial sub-populations.^9^ Most affinity-based isolation techniques, however, lend themselves more to genotypic methods (such as qPCR) as opposed to microscopy-based methods, mainly due to the use of beads which can preclude microscopic observation due to substantial loss of spatial information and loss of population heterogeneity information from the sample.

In contrast, microfluidic platforms have been increasingly used to enable bacterial enrichment compatible with microscopy. These platforms typically employ size-selection modules in the form of hydrodynamic traps in order to capture or slow down bacterial cells so as to render them quasi-static for microscopic investigation.^5,10,11^ A common issue associated with conventional bacterial size-based capture platforms are the sub-micron feature dimensions required to achieve bacterial capture. Sub-micron channel regions result in very high hydrodynamic resistances which, given the ∼5 bar burst pressure of conventional polydimethylsiloxane (PDMS)-glass devices, result in operational flow rates of only a few µLs per minute, which in turn drastically reduce the maximum sample processing speed.^12^ Although this limitation does not affect bacterial capture significantly (as parallel channels can be incorporated in microfluidic architectures to increase the speed at which the biofluid sample can be processed), it does limit downstream cell assays requiring multiple reagent infusions, e.g., sequential fluorescence *in-situ* hybridization (FISH) techniques such as multiplexed error-robust fluorescence *in-situ* hybridization (MERFISH).^13^ Identifying species using FISH by targetting hypervariable regions of the 16S-rRNA is a common rapid technique to ascertain the identity of bacterial species down to the strain level. In recent years, 16S-rRNA FISH techniques have also become more scalable via barcoding strategies (akin to those in MERFISH), an example of which is high phylogenetic resolution microbiome mapping by fluorescence *in-situ* hybridization (HiPRFISH)^14^, which enables the assay of panels containing thousand of bacterial species.

Barcoded FISH techniques are particularly attractive for the fluorescence-based identification of messenger RNA (mRNA) in spatial gene expression studies and of ribosomal RNA (rRNA) in bacterial identification studies. These techniques address the limitation of target scalability as the panel of identifiable targets may scale combinatorially with respect to the number of reagent infusion rounds performed. Shi et al.^14^ and Kandavalli et al.^15^ overcame the aforementioned sample processing speed issue by performing a spectral analogue of these techniques by using multiple complementary imager DNA strands bearing different fluorophores. For the latter study, this enabled the dye-conjugated oligonucleotide strands attached to the 16S ribosomal subunit of collected bacteria to form a spectral barcode and thus enabled combinatorial-FISH of up to 4 different imager probes leading to the identification of 7 species while minimizing the number of infusion reagent rounds. Scaling of the bacterial assay panel beyond 10 pathogens, however, would nonetheless necessitate more infusion reagents and would therefore slow down bacterial identification on this platform as a result of long reagent exchange times.^15^

Here, we achieve bacterial enrichment and identification by multiplexed 16S rRNA-FISH in a streamlined pipeline (**fig. 1**) by employing an adaptive channel to capture bacteria. This is achieved by robotic control and tuning of the channel backpressure using an in-line pressure sensor and an active feedback control system. Our approach enables the controlled formation of a region within the flow-channel with highly reduced dimensions which we refer to as the ‘capture region’. The capture region acts as a bacterial filtration element that can be formed or removed on-demand. Once bacteria are captured, the adaptive channel can be restored back to dimensions that enable high flow-rate infusion of reagents, making such platforms ideal for sequential assays. We prepare our devices using 3D-printed moulds, thus circumventing the need for complex and expensive fabrication techniques that would otherwise be needed to achieve sub-micron features. Finally, we demonstrate the identification of a panel of 7 species by evaluating simulated samples containing isolates cultured from clinical samples in the order of one-hour by using our device in tandem with multiplexed 16S rRNA-FISH. Using relative fluorescence intensity measurements for bacterial species classification, we show that our technique achieves high accuracy (in most cases, >90%) in species identification. In addition to the technique being rapid and scalable, we demonstrate that by identifying bacteria at the single-cell level, we can also discern the presence of mixed infections. Our work paves the way for using such fluidic devices for bacterial enrichment and identification in complex clinical specimens.

**Figure 1:**
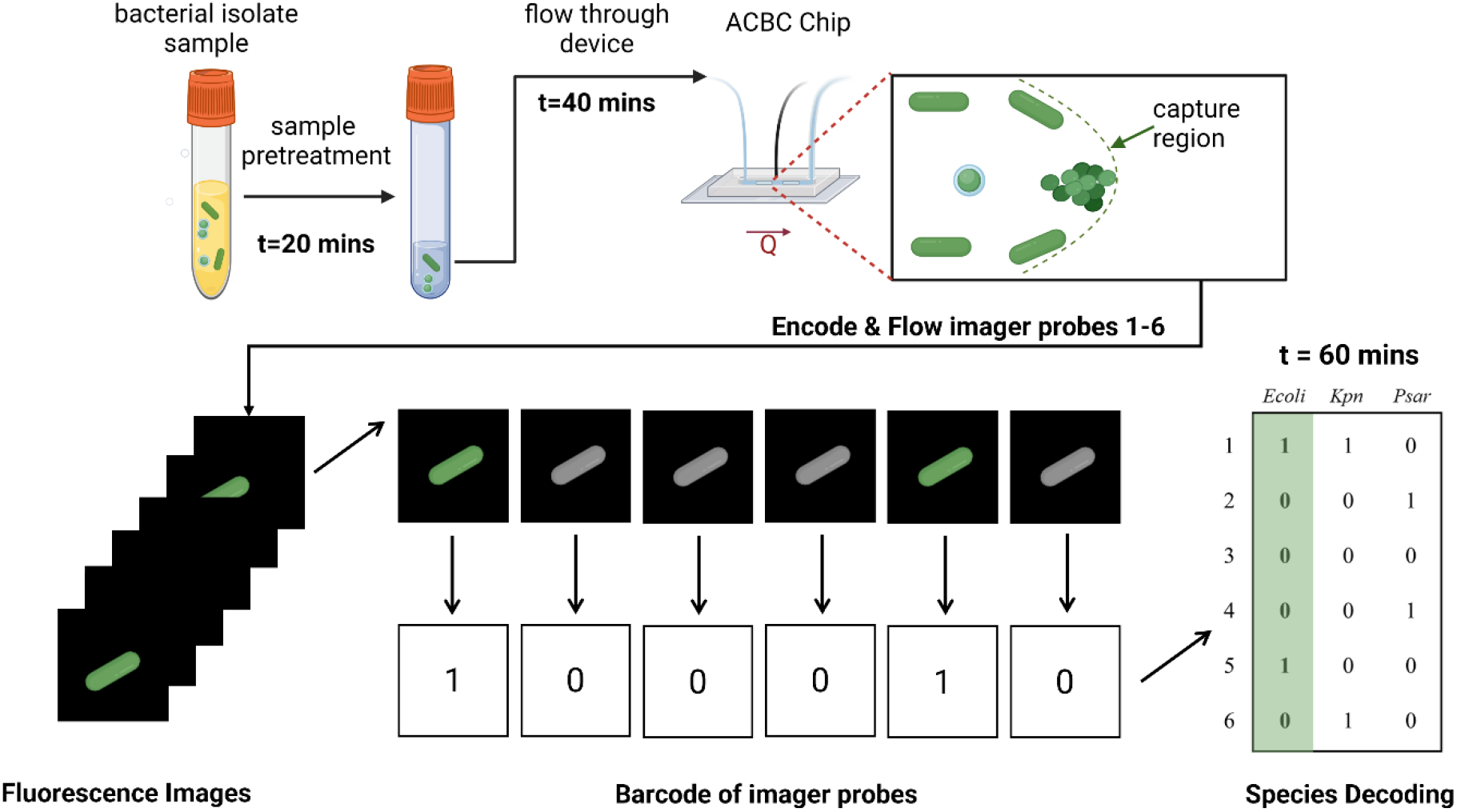
Illustration of the workflow for the bacterial capture and identification process from bacterial samples. Using the ACBC device, bacterial isolate sample is flown and captured using dynamically formed capture regions. The immobilised bacteria are then stained and assayed using multiplexed 16S rRNA FISH. The single-cell intensity of the imager probes is binarized so as to form a barcode which is then compared against a species dictionary to yield the identity of the species.

## Results

### ACBC Chip Design & Operational Principles

To address the limitations of previous approaches (especially the low operational flow-rates associated with sub-micron flow channel dimensions) while still being able to achieve bacterial enrichment, we reasoned that active formation and removal of bacterial-capture regions would facilitate both enrichment and rapid infusion of reagents after the initial capture stage of the identification pipeline.

We also designed our Adaptive Channel Bacterial Capture (ACBC) chip to accommodate the enrichment of samples containing a range of bacterial concentrations (10^2^ – 10^4^ cells/mL, with 10^2^ cells/mL reflecting patient specimens with relatively low CFU counts). We thus designed a device that captured bacteria with near 100% efficiency and immobilized bacteria to a small number of fields of view. To achieve that, we designed a bilayer PDMS microfluidic device comprising of a top control layer and a bottom fluidic layer, akin to a conventional top-down Quake valve (**fig. S1**).^16^ To achieve a pre-defined location of the capture region that was invariable to user alignment errors of the control layer, we utilized an LCD-stereolithography 3D printer so as to produce channels with both parabolic and rectangular cross-sections present on the same mould (**fig. S2**). In doing so, the capture region (**fig. 2**) was defined physically by the intersection of the channels designed with rectangular and parabolic cross-sections rather than the edge of the control channel itself, which would be prone to miniscule placement errors. We found the minimal channel width we were able to produce consistently using this approach as measured by bright field microscopy to be 34.6 ± 1.6 µm (n = 3 moulds).

**Figure 2:**
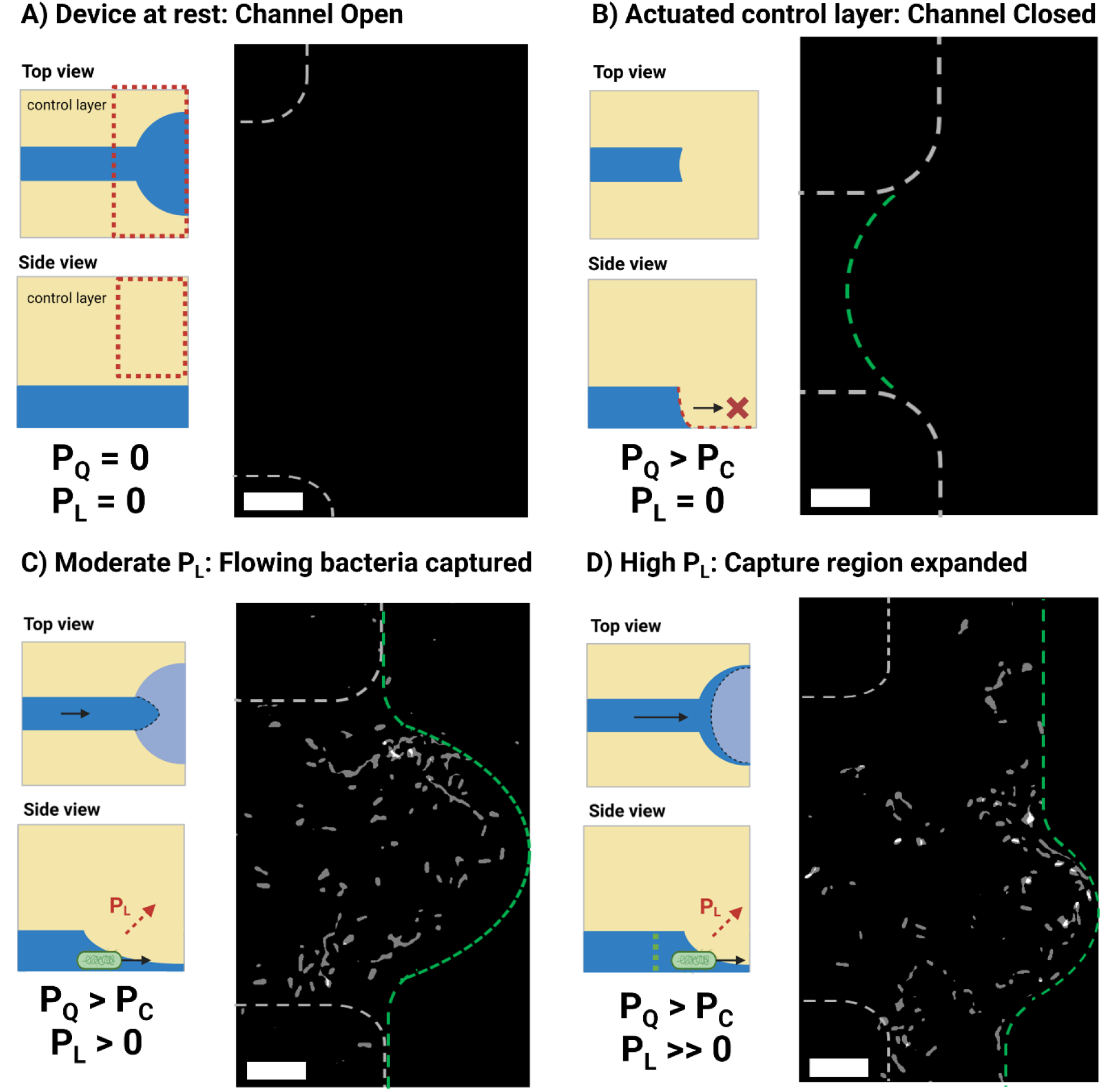
Device operation under different conditions of applied pressure **P_Q_** and lift pressure **P_L_**. Each set of conditions is depicted by a schematic as well as a position-map that showcases the end-position of *E. coli* cells flown from a PBS spike sample containing 7 × 10^2^ *E. coli/*mL across 2 runs. Dashed lines serve as a guide to the eye for the channel dimensions (white) and capture-region (green). **a)** The device at rest has its original design dimensions, no capture region is formed and thus no bacteria are trapped. **b)** The control layer is actuated at a pressure above the closing pressure **P_C_**. The channel dimensions collapse and no sample flow is observed. **c)** When a sufficient lift pressure is applied, flow through the device is enabled and a capture-region with sub-micron dimensions is formed allowing the trapping of bacteria. The end-position of flowing cells, as can be seen in the corresponding position-map, is constrained within this region. The device’s chitosan coating enables the immobilization of cells within this narrow passage. **d)** At high device backpressure values of **P_L_** the capture region can be expanded before it finally leaks across the two sides of the channel. The experimentally obtained images used to construct these maps can be found in the ESI. Scale bars correspond to 10 µm.

To perform bacterial captures, the pneumatically actuated control layer acts on a parabolic flow-channel resulting in an initially fully-sealed channel (**Fig. 2b**). Deviating from conventional Quake valve principles, and by infusing sample through the device (**Fig. 2c**) at pressures exceeding the closing pressure of the valve, the compressed fluidic channel eventually opens due to deformation of PDMS from the backpressure build-up inside the flow channel. The opening of the channel is controlled using a closed feedback-loop that maintains the channel backpressure at the desired value. When we flow bacterial cells to the capture region of our devices, they are hydrodynamically trapped at the capture-region and finally immobilized locally on the polycationic chitosan coating of the glass substrate; the immobilisation to the surface is stable, allowing the ceiling to be retracted back to full channel scale without significant release of the captured cells. By restoring the channel dimensions post-capture, we circumvent the issue that common sub-micron structured devices have with regards to operational flow rates and thus enable rapid delivery of reagents up to 50 µL per minute, drastically reducing reagent delivery times.

When attempting to use top-down valves as sieving elements in elastomer systems, it is important to note that the geometry of the microfluidic channel can alter as a function of channel backpressure due to the elastic nature of PDMS (Suppl **Video 1**).^17^ It is therefore not advised to operate syringe drivers under constant volumetric flow-rate when trying to achieve a static channel geometry, as would be necessary for the prolonged filtration of bacteria. Expansion of the channels due to pressure build-up would have the effect of releasing the captured bacteria over time. To achieve a quasi-static geometry, the syringe pumps used here operate under constant pressure control over the 20-minute infusion of the sample. Videos of PDMS deformation in the absence and presence of pressure control can be found in the electronic supplementary information (ESI) (**Videos 1-2**).

To form a channel with suitable dimensions for bacterial filtration it is important to pressurize the control channel at a pressure **P_Q_** which is above the closing pressure of the valve (**P_C_**). The **P_C_** value can be experimentally determined by pressurizing the control layer while the flow channel is at rest and visually examining the point at which bright-field contrast between the flow channel boundary and the PDMS walls (**fig. S3b**) vanishes. We found **P_C_** to be 1.3 ± 0.2 bar (n = 5 devices) for control layers fabricated using our method. This value exceeds devices made with soft lithography, which we attribute to the larger depth of our channels, as well as their partial deviations from semi-circularity. Compera et al.^18^ have shown that closing pressures of 0.25-1.3 bar were observed when using channel depths of 100 µm from 3D printed moulds which were re-flown or anti-aliased respectively, the latter of which are like the ones employed here.^18^

Using fluorescence measurements under no net-flow (**P_chip_** = 0 bar) we estimated the channel’s resulting height at different values of **P_Q_** (**fig. S3a**) and observed a linear relationship between applied pressure **P_Q_** and the resulting channel height, **h**, with a regression coefficient of 16.6 µm/bar (r^2^ = 0.98).

By infusing the flow channel with a syringe pump able to generate sufficient torque to exceed the difference of the control pressure **P_Q_** and the device closing pressure **P_C_**, an overall lifting pressure can be generated that raises the collapsed channel. We calculate this lift pressure, **P_L_**, experimentally from our device’s in-line pressure sensors using the following equation:

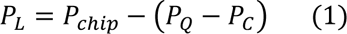

The flow-channel pressure, **P_chip_**, is calculated live and used to estimate the height of the collapsed channel of the capture region. We estimated the channel’s resulting height using fluorescence measurements at actuated control-layer conditions (**P_Q_** = 2 bar) for different values of **P_chip_** by applying flow-induced pressure to the flow layer (**fig. S3b**) and observed a linear dependence of resulting channel height, **h**, and the applied pressure. A regression coefficient of +8.6 µm/bar (r^2^ = 0.98) was determined which was lower than the magnitude of the membrane displacement coefficient observed when flow was at rest (-16.6 µm/bar). Given that PDMS exhibits linear elastic behaviour for strains up to 20%,^19^ which would not be exceeded with a membrane displacement of 20 µm (membrane thickness: 200 µm), we presume that this difference is associated with the direction of pressure as applied by the flow-channel to the PDMS membrane. Indeed, when we pressurize the flow-channel, in addition to changes in channel height we also observe receding of the membrane along the flow-path direction. This suggests that stress is distributed both upwards but also longitudinally to the flow-path direction and could explain the difference in the regression coefficients observed in the two scenarios.

Under our operational conditions (**P_Q_** = 2 bar, **P_chip_** = 1 bar, **P_L_** = 0.3 bar), **h** was estimated to be 2.3 µm at the edge of the capture region. This estimate is larger than the typical diameter of bacteria, however in practice we observe near 100% capture efficiency of bacteria at these conditions. At the above conditions the stress along the flow-channel dimensions results in the planar projection of the capture region to be parabolic in shape with maximum displacement in the order of 25 µm as can be seen below (**fig. 2C**).

We found that operating at a pressure **P_Q_** of 2 bar or higher (which is >>**P_C_**) is more practical, since operating at **P_Q_**=**P_C_** led to more pronounced effect of backpressure variations on the final geometry of the capture region. Further, a **P_Q_** of 2 bar did not damage the PDMS membrane over prolonged periods of actuation. The boundary of the capture region can be visually observed using bright field microscopy as well as by the autofluorescence of PDMS when excited at short wavelengths (405-473 nm) in highly inclined thin illumination (HILO) mode (**fig. S4**). This is a useful feature as it enables placement tuning of the capture region as desired depending on the experiment at hand. For instance, we found that for samples with high bacterial densities (10^7^ bacteria/ml) that the narrow region of our device would result in very dense packing of bacteria over time, in which case the capture region area could be expanded towards the wider section of the device by allowing the backpressure of the device to build up further (**fig. 2d**). This in turn allowed for much more uniform spreading of the cells on the chitosan surface, simplifying bacterial imaging of these samples.

### Bacterial Capture and Recovery

To assess the device’s performance at capturing bacteria at low cell densities, we performed capture experiments with *E. coli* spiked at a known concentration into phosphate buffer saline (PBS) and subsequently flowing the solution through our device over the course of 20-minute runs. This was performed by monitoring the autofluorescence of *E. coli* grown in Luria-Bertani (LB) broth after excitation using a 473 nm laser. Monitoring the trajectory of flowing bacteria using this method enabled both visualization of the trajectory of incoming cells and counting of the number of cells able to pass beyond the capture region. Using combinations of **P_Q_** and **P_chip_** such that a capture region is formed but not over-expanded (**P_Q_** = 2 bar, **P_chip_** = 0.8-1.1 bar), we achieved ∼100% capture efficiency (see below) of flowing cells over the course of 20-minute flow-through experiments. (**fig. 3b**).

**Figure 3:**
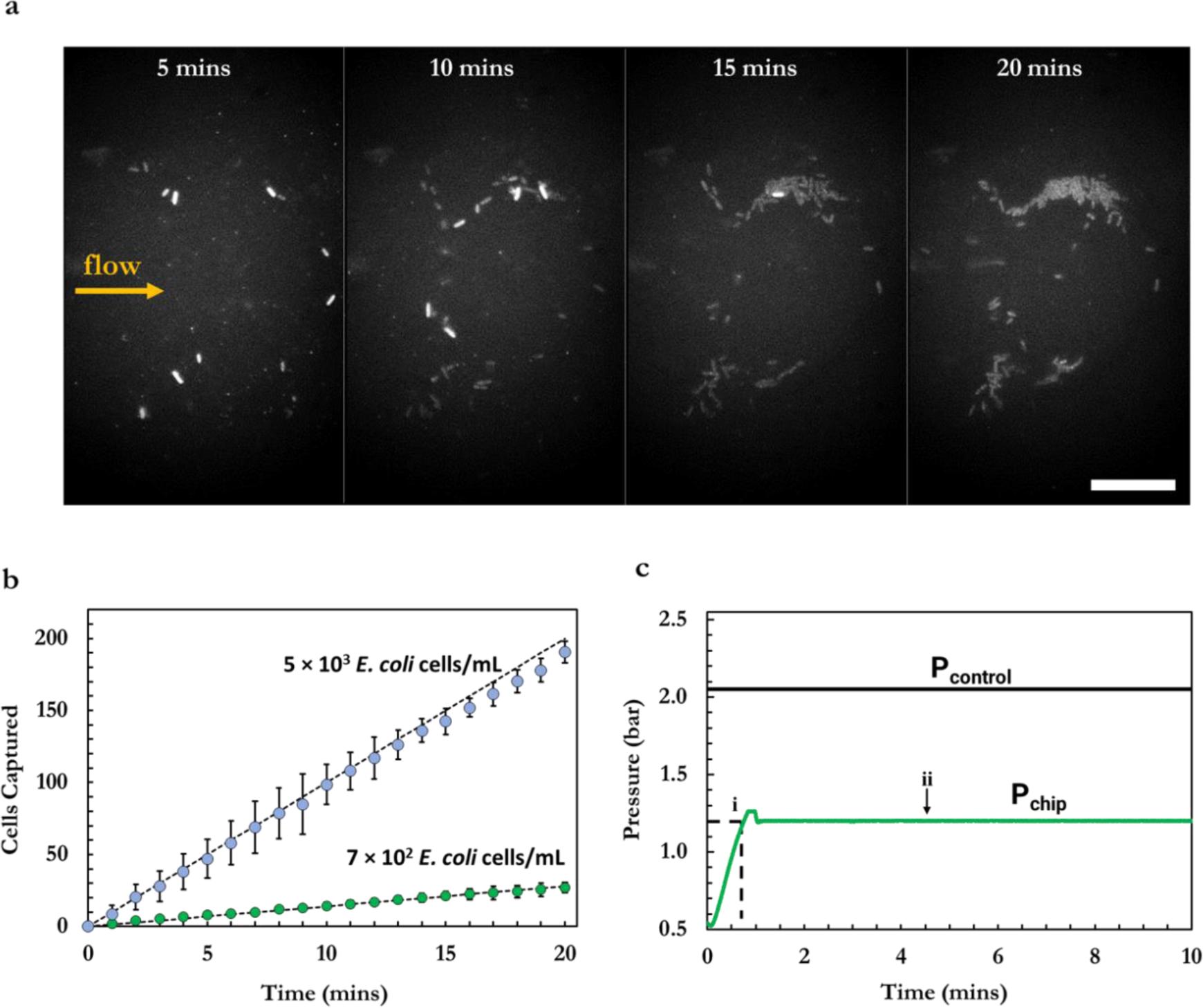
**a)** Autofluorescence timelapse of MG1655 *E. coli* cells collected in the ACBC capture region from a 7 × 10^2^ *E. coli/*mL sample over 20 minutes. Newly arriving cells display higher autofluorescence intensity relative to cells imaged over the course of the timelapse due to photobleaching of the innate cell fluorescence over prolonged exposure. Scale bar is 15 µm. **b)** Number of cells captured over the course of a 20-minute filtration for a sample with a cell count of 7 × 10^2^ *E. coli/*mL (n = 4) and 5 × 10^3^ *E. coli/*mL (n = 3). On average, 27 and 191 *E. coli* cells were captured by the end of 20-minute course runs respectively. The dashed black lines denote the theoretical maximum capture from samples at these concentrations. **c)** Exemplar pressure trace obtained during capture runs. With the adaptive ceiling actuated, sample is infused through the flow-layer resulting in an initial increase of **P_chip_**. When capture region formation is observed **(i)**, the syringe pump is instructed to maintain this value of backpressure. Initial overshooting to the target pressure can be seen before stabilization of the backpressure to the desired value **(ii)**.

The maximum flow-rate through the device while producing backpressures able to sustain a capture region was found to be 2.1 ± 0.3 µL/min. For dilute samples containing 7 × 10^2^ cells/mL, 20-minute flow-through runs were found to yield 26.8 ± 3.1 captured bacterial cells (n=4). This number of cells captured is within error from the average value of 29 cells expected to be captured at 100% capture efficiency. At this processed sample volume (42 µL) and sample concentration, stochastic sampling effects are not expected to be significant and therefore the expectation value of 29 cells of sample at these conditions is a reasonable estimate. Furthermore, no fluorescence trajectories of cells bypassing the capture region were observed during flow-through acquisitions. These findings therefore suggest that there are only negligible losses of cells through the device and that the capture region is capable of achieving bacterial filtering at 100% capture efficiency.

When the adaptive ceilings of the device were not actuated (e.g., conditions as in **Fig. 2a)**, we did not observe the capture of any bacteria within the capture region observation field suggesting that the chitosan coating alone, used to immobilize bacteria post-capture, is not sufficient to isolate an appreciable number of cells at this cell density. Figure 2 denotes the end position of *E. coli* bacteria captured across a series of runs for fully sealed capture regions (n=2) and expanded capture regions (n=2). As seen in the fully sealed case (**fig. 2c**), the end-position of bacteria is localized within a narrow region and bacteria are attached uniformly within this area. When higher **P_chip_** pressure is applied, in addition to the height increase of the membrane, the boundaries of the capture region are further displaced downstream (16 μm displacement across the boundary), effectively expanding the capture region. Notably, when no chitosan coating was used, the end positions of bacteria were observed predominantly at the boundaries of the capture region (Suppl **Video 2** and **Video 3**).

It was important to ensure that the ACBC device was able to capture a wide range of bacterial species to ensure that a variety of bacterial pathogens could be identified using this platform. Hydrodynamic trap formats act as low-pass filters and therefore the capture region of the ACBC device could potentially be unable to trap flowing bacteria of smaller dimensions. To verify the capabilities of the device to capture smaller diameter bacterial species we performed capture runs of a *S. aureus* (d = 0.5 – 1.5 μm) strain (**Video 4**).^20^ We found that *S. aureus* cells are captured efficiently, suggesting that the ACBC chip is capable of capturing smaller diameter bacteria as well. A capture timelapse for *S. aureus* captures can be found in the supplementary information (**Video 4**).

Following microscopic investigation, trapped bacteria could be released from the chitosan coated surface of our device by flowing alkaline lysis buffer (0.2 M NaOH, 1% SDS) through the device. This could potentially enable further downstream analysis of the sampled cells to be carried out, e.g., for sequencing methods (**Video 5**).

### Multiplexed 16S rRNA-FISH for Bacterial Identification

To demonstrate that the device can support processing and further analysis of the captured bacteria, we developed methods to identify a panel of seven pathogenic bacteria commonly associated with human infections (**fig. 4**). These species contribute to many cases of serious human infection, including bloodstream and joint infections, and meningitis.

**Figure 4:**
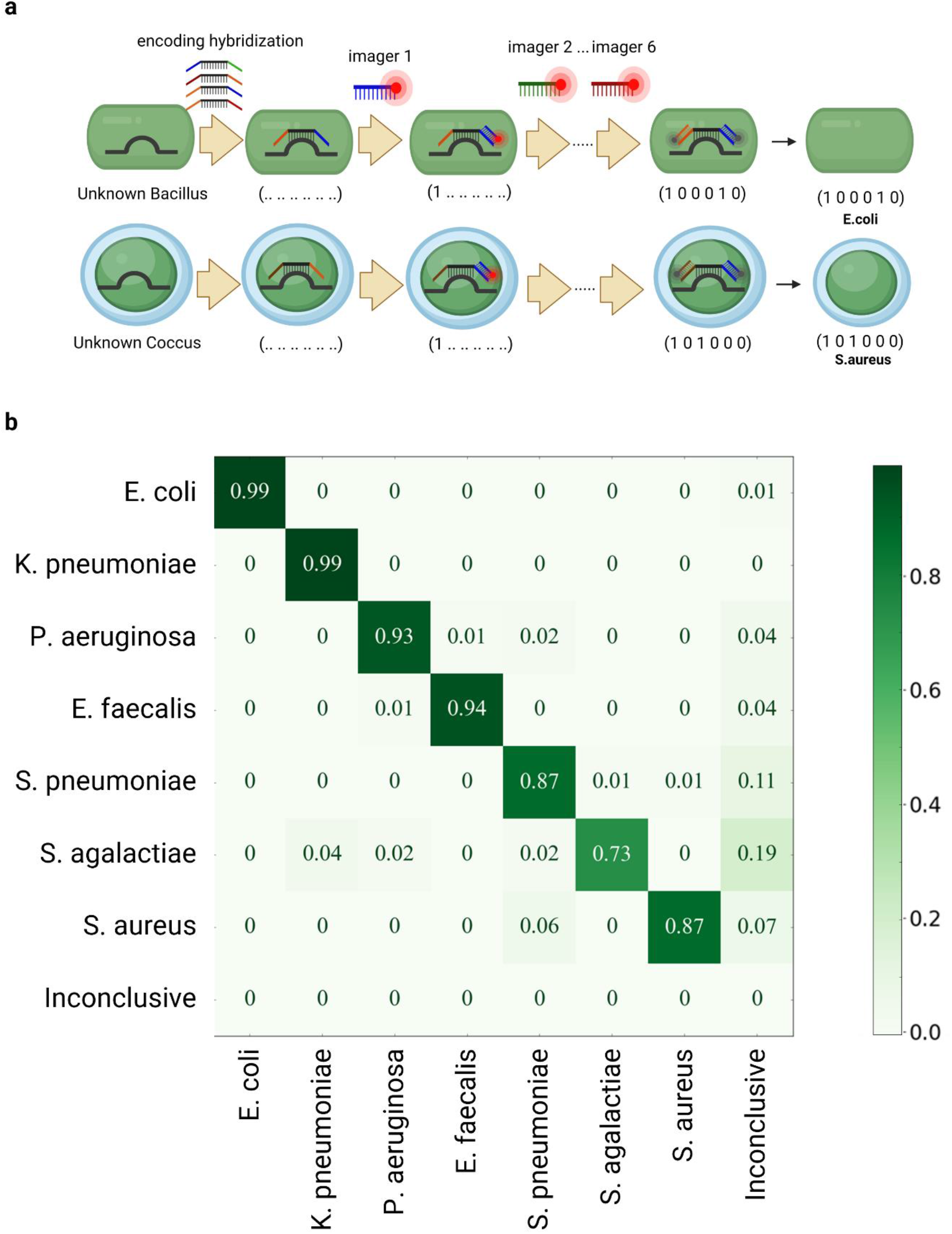
**a)** Schematic of the multiplexed 16S rRNA assay. The rRNA of the captured bacteria is hybridized with encoding probes containing two landing pads. Six rounds of hybridizations with fluorescently tagged oligos then reveal the identity of the bacterial species. **b)** Confusion matrix of this binary classification method showing classification rates for all species assayed from bacterial isolates. The inconclusive class corresponds to any barcode raised that does not correspond to an entry in the species dictionary.

Specifically, we employed encoding oligonucleotides containing a 30-nucleotide (nt) targetting sequence, flanked by two 25-nt “coding sequences”. To design these sequences, 16S rRNA targetting sequences were first identified from literature studies that deployed these probes in human samples (**table S1**); this was done to ensure there was no cross-interference between the probe and common host sequences present in clinical sample matrices. These targetting sequences were aligned against strains found in the NCBI’s database of 16S ribosomal RNA sequences for bacteria and archaea^21^ and extended to 30-nt to ensure all sequences exhibited similar melting temperatures and more resiliency towards stringent probe washes. The targetting sequences were finally flanked on both sides by 25-nt coding sequences obtained from the Elledge group’s database of orthogonal oligos^22^ while ensuring the assembled oligo showed no continuous alignment beyond 14 base pairs to either the 16S targetting sequences nor the genome of the panel species.

To perform the assay on captured bacteria, we performed an initial 20-min hybridization step containing all encoding probes, and then washed all weakly bound probes. The coding sequences were subsequently hybridized and imaged using complementary fluorescent Cy5-imager probes which were flown sequentially through the ACBC chip (**fig.4a**). The performance of each infusion round was assessed by monitoring the integrated single-cell fluorescence intensity resulting from binding of the imager probes, and the identity of the species was decoded from a species dictionary after six imaging rounds.

To assess the performance of our assay in identifying bacterial species, we employed a classifier which first normalized the single-cell intensity signal of the 6 imager probes based on the intensity of the brightest probe, and identified the two imager probes exhibiting the highest single-cell intensity. All other probe signals were regarded as null. If either of the two highest single-cell signals obtained were below a threshold value of 2 standard deviations above non-specific background, then they were also regarded as a negative (i.e., a “0” entry), and the species was subsequently classified as inconclusive. After converting the 6-imager signals to a binary six-entry vector barcode, this barcode was cross-referenced against a dictionary collated from the assigned bacterial barcodes termed here as the “species dictionary”. In addition, if a barcode was raised that was not present in this dictionary, then the cell was designated to the ‘inconclusive’ class suggesting that no identification could be made. Otherwise, one of the 7 species classes was assigned.

Our unsupervised approach was able to achieve high classification accuracies at the single-cell level, which we have categorized into three classes.The first class, which we refer to as ‘very high accuracy species’, includes *E. coli* and *Klebsiella pneumoniae*, both with a classification rate of 99%, and *Pseudomonas aeruginosa* and *Enterococcus faecalis*, with rates of 93% and 94% respectively. The second class, termed ‘high accuracy species’, consists of *Streptococcus pneumoniae* and *Staphylococcus aureus*, both achieving a classification rate of 87%. The third and final class, the ‘moderate accuracy species’, is represented by *Streptococcus agalactiae*, which had a classification rate of 73% (**fig 4b**).

This variation in classification accuracy of the assay across the three classes of bacteria can be attributed, at least in part, to the differential susceptibility of bacterial populations, particularly Gram-positive species, to lysozyme treatment. Lysozyme treatment is necesssary in order for FISH probes to access the ribosomal content. However, within a given bacterial population, individual bacteria may exhibit varying degrees of susceptibility to lysozyme treatment over the 20-minute treatment period (**fig. S5b**). This variability in permeabilization can impact the accessibility of the probes to the ribosomal content of the more lysozyme resistant cells, which in turn can affect the classification accuracy. Specifically, species that are less susceptible to lysozyme treatment, and therefore less permeabilized, are likely to fall into the ‘moderate’ and ‘high’ accuracy classes.

While higher lysozyme concentrations could enable both faster accessibility and larger fraction of permeabilized ‘high’ and ‘moderate’ accuracy class bacteria, we observed that the remaining bacteria in this panel can lyse at higher lysozyme conditions. As a result, and considering the excellent performance of the classification assay across all species, this set of conditions was chosen for its ability to universally allow classification across all species in the present panel.

Beyond the multiplexed 16S rRNA assay that facilitates bacterial species identification, we also employ a pan-bacterial probe, EUB338-Cy5 (**fig. 5a**). This probe serves a dual purpose: it detects the presence of bacteria not specifically targeted in the probe panel, and aids in segmenting and distinguishing bacteria from matrix components. The detection of the presence of bacteria in a biological sample is a critical clinical finding in itself, underscoring the value of this comprehensive approach.

**Figure 5:**
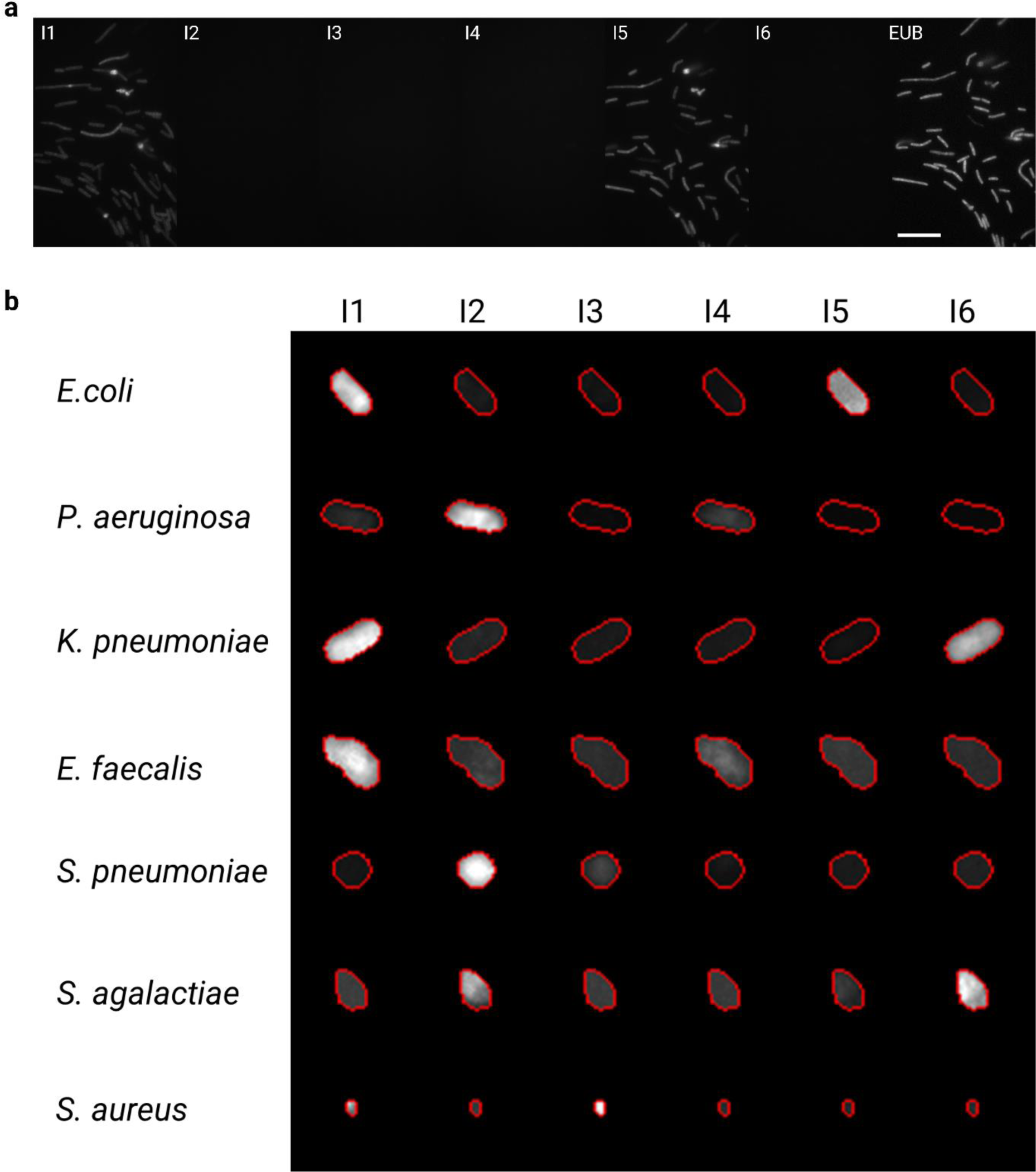
**a)** Exemplar sequential hybridization images of an *E. coli* bacterial strain assayed on the ACBC device. We note that a small number of bacteria detach from the chitosan coating of the device over the course of the sequential hybridization infusions. Scale bar is 15 µm. **b)** Exemplar single-cell bacteria images and their relative single-cell fluorescence intensities (normalised to brightest probe) after assay with the multiplexed 16S-rRNA imager probes.

Of course, when guiding a diagnosis, a decision would not be made on the basis of a single-cell classification alone. This is due to the inherent variability among individual cells within a population, which can lead to discrepancies when extrapolating single-cell data to a broader context. Therefore, the potential for correct classification of an infection is likely to be increased when operating at the sample level, as this approach takes into account the broader context of the bacterial population (i.e. facilitates bacterial presence/absence assessment based on imaging multiple rather than individual bacterial cells).

An advantage of having single-cell based resolution for identification is in the ability to detect the presence of multiple pathogens in an infection. The ability to differentiate between species in mixed infections is particularly crucial in clinical settings, where accurate identification can guide targeted antibiotic treatment strategies. Mixed infections often require a more nuanced approach to treatment compared to mono-bacterial infections, as they may involve pathogens with different antibiotic resistance profiles.

To demonstrate that we can distinguish mixed infections using our multiplexed 16S rRNA-FISH assay such as in the case of the two Gram-negative rods *K. pneumoniae* and *P. aeruginosa*, we stained the former with a membrane stain, Wheat Germ Agglutinin (WGA-AF488), and immobilized the two species to a chitosan-coated surface mixed at a 1:1 cell density ratio. After immobilization and after noting the position and identity of the bacteria, the species were treated with permeabilization solution and hybridized with the encoding probe solution. The identity of the two species was finally verified using the sequential imager probe hybridizations.

These two species would have been hard to distinguish by purely morphological features or Gram-stain differentiation (**fig.6b**).; the precise identification and differentiation of pathogens within a mixed infection, as enabled by our assay, can inform the selection of more effective, personalized antibiotic regimens.

## Materials and Methods

### Preparation of ACBC Devices

Flow and control channel moulds of microfluidic devices were designed using Autodesk Fusion360 and printed with an LCD stereolithography 3D printer (Anycubic Mono 4k, Shenzhen, China) using Anycubic ECO plant-based clear resin. The moulds were washed with IPA, further UV cured for 15 minutes and placed in an oven at 70°C for two days to eliminate uncured resin residues that have otherwise been found to inhibit PDMS crosslinking.

For the flow-layer, PDMS (Sylgard 184, Dow Corning, United States) mixed at a ratio of 20:1 was spin-coated (Ossila spin-coater, Ossila ltd, The Netherlands) at 500 RPM on the flow-layer mould. For the control layer, PDMS was mixed at a ratio of 5:1 on a 3D printed mould until 2 mm the height of PDMS reached 2 mm. The two moulds were then placed in an oven at 70°C for 35 minutes. The off-ratio bonding technique was then employed (**fig. 7**). Briefly, the moulds were removed from the oven and allowed to cool down. The cured control layer PDMS negatives were then cut out, and inlets were punched using a biopsy punch (1 mm) and aligned to the flow-layer devices under a microscope. PDMS mixed at a ratio of 5:1 was then poured around the control layer to form a device with a final thickness of 2 mm and cured in an oven at 70°C for 1 hour. The PDMS device was finally detached from its mould, inlets were punched using a biopsy punch (1 mm) and finally bonded to glass after the two parts were treated with air plasma. Devices were placed in an oven at 70°C overnight to further reinforce bonding. The devices were finally interfaced using steel pins (gauge: 5G) fitted to silicone tubing (Tygon, ID: 250 µm ID). For medium pressure interconnects, the interconnection was reinforced by using epoxy (Araldite fast-cure) at the steel-PDMS interface, allowing the epoxy to harden for 6 hours and finally an additional baking step at 70°C for 2 hours.

**Figure 6:**
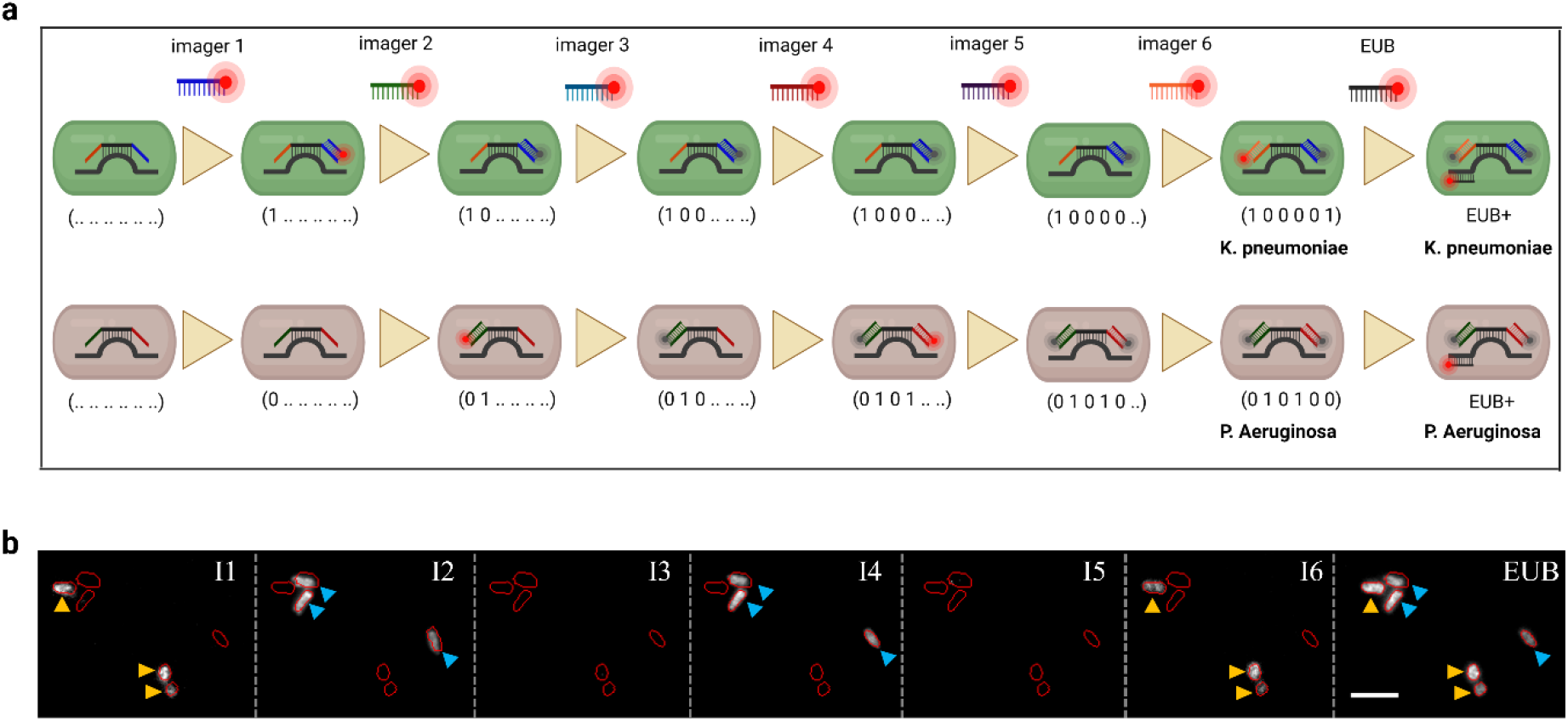
**a)** Schematic showcasing a mixed infection scenario. **b)** A field of view showcasing a mixed infection scenario by the gram-negative rods *K. pneumoniae* and *P. aeruginosa*. True-positive imager probes for the former (**I1** and **I6**) and the latter (**I2** and **I4**) selectively bind to their respective species target suggesting minimal cross-binding of the encoding probes. Negative probes (**I3** and **I5**) show minimal fluorescence signal above background upon excitation. The pan-bacterial kingdom probe, EUB338, is employed at the end of the assay to stain all bacteria present in the field of view. Scale bar is 5 µm.

**Figure 7:**
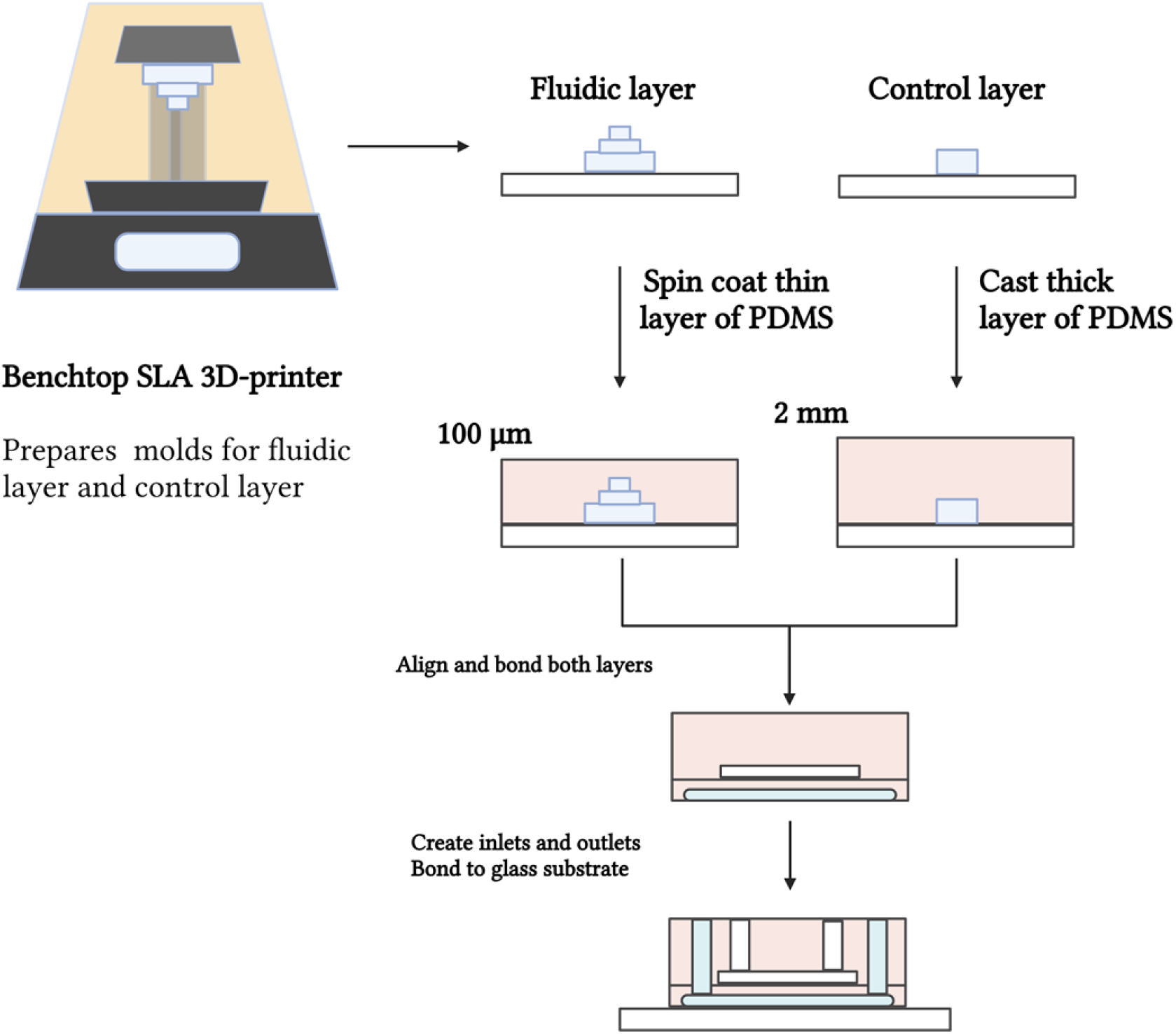
Schematic showing the benchtop preparation of ACBC devices from 3D printed moulds. Both fluidic and control layers are printed, cast with a thin and a thick layer of PDMS respectively and finally aligned and bonded so as to form the final device architecture.

### Flow instrumentation

The fluidic setup consisted of a syringe pump (Elite 11, Harvard Apparatus, USA) operated in constant pressure mode used to infuse the sample as well as 8 custom-made Arduino controlled syringe pumps used to deliver assay reagents via a Labsmith 8-port selector valve (AV801) (**fig. S6**). A 3-way selector valve (Labsmith, AV201) is connected to the main chip line and selects whether the sample containing syringe pump or the assay reagent valve are engaged for infusion to the chip. Prior to the ACBC device inlet, an in-line pressure sensor (LS-uPS0800) is placed to measure the ACBC device backpressure P_chip_. For adaptive channel control, a variable constant air pressure source was connected to an in-line pressure sensor (LS-uPS0800) and interfaced to the control layer of the ACBC device.

### Bacterial strains and preparation of simulated bacterial cultures/mixtures

Single, banked clinical isolates of *E. coli, P. aeruginosa, K. pneumoniae* (Gram-negative) and *E. faecalis, S. pneumoniae, S. agalactiae*, and *S. aureus* (Gram-positive) were grown in Brain Heart Infusion (BHI) broth supplemented with 2% yeast extract overnight at 37°C and subsequently diluted to OD_600nm_ in fresh broth for further growth until an OD_600nm_ of 0.2 was reached. Bacterial cells were then fixed in a PBS solution containing 2% paraformaldehyde for 20 min. Cells were then centrifuged and the pellet resuspended and permeabilized in a 1:1 Ethanol/PBS solution and stored at -20°C until use.

### Preparation of bacterial samples for capture studies

Preliminary bacterial capture studies were conducted with MG1655 *Escherichia coli*, a lab-adapted non-pathogenic K-12 derivative. MG1655 cells were grown in LB broth until an OD_600nm_ of 0.2 was reached. Bacterial cells were then fixed in a PBS solution containing 2% paraformaldehyde for 20 min. Cells were then centrifuged and the pellet resuspended and permeabilized in a 1:1 Ethanol/PBS solution and stored at -20°C until use. For exemplar ACBC capture videos (Suppl **Video 1** and **Video 2**), the same protocol was followed, except the cells were also stained with Nile red membrane stain (15 minute incubation, 1 μg/mL) for cell circumference visualization.

### Bacterial capture using ACBC device

For capture efficiency studies, devices were first surface-treated with Chitosan (0.015% w/w in 0.1 M acetic acid) for 10 minutes and then washed with 1X PBS. Fixed MG1655 *E. coli* cells grown in LB were flown and enumerated using their native autofluorescence resulting arising from the rich growth medium (473 nm excitation). The densities of all cell suspensions were determined using a Neubauer haemocytometer.

The ACBC control layer, filled with milli-Q water, was then actuated at 2 bar, after which bacteria containing sample was flown through the device at a flow rate of 40 µL min^-1^. The backpressure of the system was monitored throughout. The formation of a capture region was observed visually as backpressure built up within the device and the syringe pump controlled via custom software was then instructed to maintain constant pressure so as to not perturb the geometry of the capture-region. The collection of bacteria was then allowed to proceed for 20 minutes, or until a sufficient amount of target was deemed to have been collected.

### Imaging

Fluorescence images were captured using a wide-field Nanoimager microscope (ONI, Oxford, UK) equipped with a Hamamatsu Flash4 v3 sCMOS camera. Samples were imaged in highly inclined thin illumination (HILO) mode using a 100× oil-immersion objective. The laser illumination was focused at 2° below TIR which was at an angle of 51.5° with respect to the normal. For multiplexed FISH assays, images were acquired at an exposure time of 33 ms using a 640 nm excitation laser at a power of 0.78 kW/cm^2^. For tracking of the bacteria during flow-through experiments, the sample was illuminated for an exposure time of 500 ms using a 473 nm laser at a power of 1.09 kW/cm^2^.

### Bacterial segmentation

For bacterial segmentation we employed Cellpose,^23^ an instance segmentation model, to segment cells from epifluorescence images of *E. coli* (MG1655) labelled with EUB338-Cy3 probes excited with 532 nm light. To improve segmentation performance, we trained a custom Cellpose model on our microscopy data for 100 epochs using the standard Cellpose hyperparameters. Cellpose segmentation and segmentation curation/editing was carried out in Napari-BacSeg, which is a custom Napari plugin that was built for segmenting and analysing images of bacteria. Napari-BacSeg is available from the Napari Hub.

### Multiplexed 16S-FISH

Fixed bacterial isolates were centrifuged at 7,000 g for 5 minutes, re-suspended in PBS and further permeabilized using a Lysozyme solution (20 mg mL^-1^) dissolved in TEG buffer (25 mM Tris, 10 mM EDTA, 50 mM Glucose, pH = 8) for 20 minutes at 37°C. The bacteria were then either immobilized on a glass surface treated with Chitosan (0.015% w/w in 0.1 M acetic acid, 10 min treatment time) for 20 minutes or captured and immobilized within an ACBC device for 20 minutes or until a sufficient amount of bacteria was collected. The permeabilized bacteria were treated with encoding hybridization buffer (1X Denhardt’s reagent, 10% Dextran Sulfate, 2X SSC, 30% formamide, 1 mg mL^-1^ yeast t-RNA, 0.04% SDS, EP ssDNA probes) for 15 minutes and then washed using encoding wash buffer (2X SSC, 30% formamide). For Gram-negative species, 1 µM EP ssDNA probes were used, whereas for Gram-positive species 5 µM of EP ssDNA probes were used. The encoding probe sequences can be found in the supplementary information (**Table S1**).

Following the encoding process, the sample was then sequentially interrogated using imager probe (I1-I6) hybridization solutions (1X Denhardt’s reagent, 10% Dextran Sulfate, 2X SSC, 10% formamide, mg mL^-1^ yeast t-RNA, 0.04% SDS, 100 nM IP-Cy5 ssDNA probe) for 1 minute, followed by washing of weakly bound probes with imager wash buffer (40% formamide, 0.2X SSC). Following fluorescence imaging of the imager probe, the fields of view of interest were subsequently photobleached prior to the introduction of new imager probes. The imager probe sequences can be found in the supplementary information (**Table S2**). For validation of bacterial presence the EUB338-Cy5 probe (**Table S2**) was used (2X SSC, 10% formamide, 1 mg mL^-1^ yeast t-RNA, 0.04% SDS, 100 nM EUB probe).

## Discussion

In this study we have demonstrated the development of a microfluidic platform capable of capturing and identifying common human bacterial pathogenic species using adaptive channels capable of forming capture regions for bacteria. By carefully controlling the channel backpressure, we show that sustained bacterial trapping can be achieved. The actuated nature of the system enables the device to act either in capture mode or interrogation mode, circumventing a major issue that common hydrodynamic trapping-based devices have limits infusion of reagents at high flow-rates. In the capture mode, our device can isolate micron-sized objects from solution, albeit at low flow-rates (2.1 µL min^-1^); in interrogation mode, however, reagents can be flown at much faster rates (up to 50 µL min^-1^) and therefore enable the rapid exchange of reagents as would be needed with current state of the art single-cell transcriptomic techniques.

Future work in the design of the ACBC chip will focus on assessing and increasing clinical sample processing speeds, which will rely on introducing parallel elements in the device design, as well as tuning the hydrophilicity of the capture-region channel that forms upon channel collapse. Further refinements in the architecture of future devices will also be investigated to mitigate the need for surface chemistry modifications required to achieve bacterial-immobilization post-capture. Such approach will enable the microscopic investigation of bacteria otherwise not as prone to immobilization in the poly-cationic surfaces presently used (chitosan) in the ACBC device. We expect these modifications to further facilitate the passage of the aqueous sample matrix, achieve higher overall flow rates under the same channel backpressure conditions and enable universal capture and assay of bacteria found in clinical samples.

Using the ACBC device, we have shown that we can capture bacterial cells from simulated samples in which patient isolates are spiked in known concentrations into buffers. We have also shown that the species of these captured bacteria can be reliably ascertained using a multiplexed 16S-rRNA FISH molecular barcoding method that achieves high classification rates for seven species in an unsupervised manner. In addition to mono-bacterial infections, our assay’s capacity to identify and differentiate multiple pathogens within a single sample can provide a more comprehensive understanding of polymicrobial infections. This could potentially reveal interactions between different bacterial species within the infection, offering further insights into the pathogenesis and progression of the disease. Ultimately, these detailed insights could contribute to the development of more effective treatment approaches for patients.

While a limited number of patient isolates were evaluated, the performance of this combined capture and identification approach yielded high bacterial species classification rates relevant to the typical CFUs of these samples. We aim to further scale the panel of pathogens that can be currently assayed although we do envisage that potential difficulties will eventually arise due to species homology overlaps in the 16S hypervariable regions as more species are included in the assay panel used. However, we remain confident that by adopting more elaborate barcoding schemes and also by introducing error correction methods in the species dictionary employed, the identification of a wider range of bacteria using this method will be realized. In addition, the inclusion of a universal bacterial probe in principle enables the detection of bacteria not present in the panel, and this result in itself could also be relevant in empiric antibiotic prescribing decisions.

## Author Contributions

S.C. designed the ACBC device, carried out experiments, analysed data and wrote the manuscript. P.T. developed software tools used for segmentation and analysis of bacteria. C.F. developed protocols for bacterial isolate culture and provided the strains used in this study. A.F. trained ribosome classifier models used for bacterial segmentation. H.E.S. and J.K. performed preliminary experiments and provided reagents. S.C., A.N.K., N.S. and C.N. conceived the study and interpreted results. All authors reviewed and approved the final manuscript.

## Acknowledgements

This work was supported by an Oxford Martin School (by the establishment of the Oxford Martin School Programme on Antimicrobial Resistance Testing; to ANK, NS, CN), by Wellcome Trust grant 110164/Z/15/Z (to ANK) and by UK Biotechnology and Biological Sciences Research Council grants BB/N018656/1 and BB/S008896/1 (to ANK) and by Oxford’s EPSRC/BBSRC Impact Acceleration Account EP/X525777/1 (award 0012379, to ANK, NS, and CN). The research was additionally supported by the National Institute for Health Research (NIHR) Health Protection Research Unit in Healthcare Associated Infections and Antimicrobial Resistance (NIHR200915) at the University of Oxford in partnership with United Kingdom Health Security Agency (UKHSA) and by the NIHR Oxford Biomedical Research Centre. This work was also supported by a John Fell Fund grant to NS (Grant application: 0008776). NS is an NIHR Oxford BRC Senior Research Fellow. Figures presented contain elements created with Biorender.com

## Ethics

Ethical approval for the use of clinical samples and isolates processed by the John Radcliffe Hospital microbiology laboratory in the development of diagnostic assays was granted by the UK’s Health Research Authority (London - Queen Square Research Ethics Committee [REC reference: 17/LO/1420]).

## Data availability statement

The data that support the findings of this study are available from the corresponding authors upon reasonable request.

## Code Availability

Code used to control fluidic hardware, generate the moulds from stereolithography files and generate the results is available from the corresponding authors upon reasonable request.

