## Supplemental Information for "Rapid Identification of Bacterial isolates Using Microfluidic Adaptive Channels and Multiplexed Fluorescence Microscopy"

**Table of contents:**

- **Figure S1:** The layout of the ACBC microfluidic device featuring all dimensions used.
- **Figure S2:** Channel profiles resulting from 3D printed moulds.
- **Figure S3**: Fluorescence measurements of adaptive channel height as a function of applied pressure.
- **Figure S4**: Visualization of the capture region.
- **Figure S5**: Fluidic setup used for ACBC chip capture of bacteria.
- **Figure S6**: Permeabilization of a pathogenic *E. coli* isolate in TEG buffer.
- **Figure S7**: Trapping of bacteria in a non-adherent device conditions.
- **Table S1**: Table containing the encoded FISH probes used for species identification.
- **Table S2**: Table containing the imager FISH probes used for species identification.

### Layout of the ACBC microfluidic device

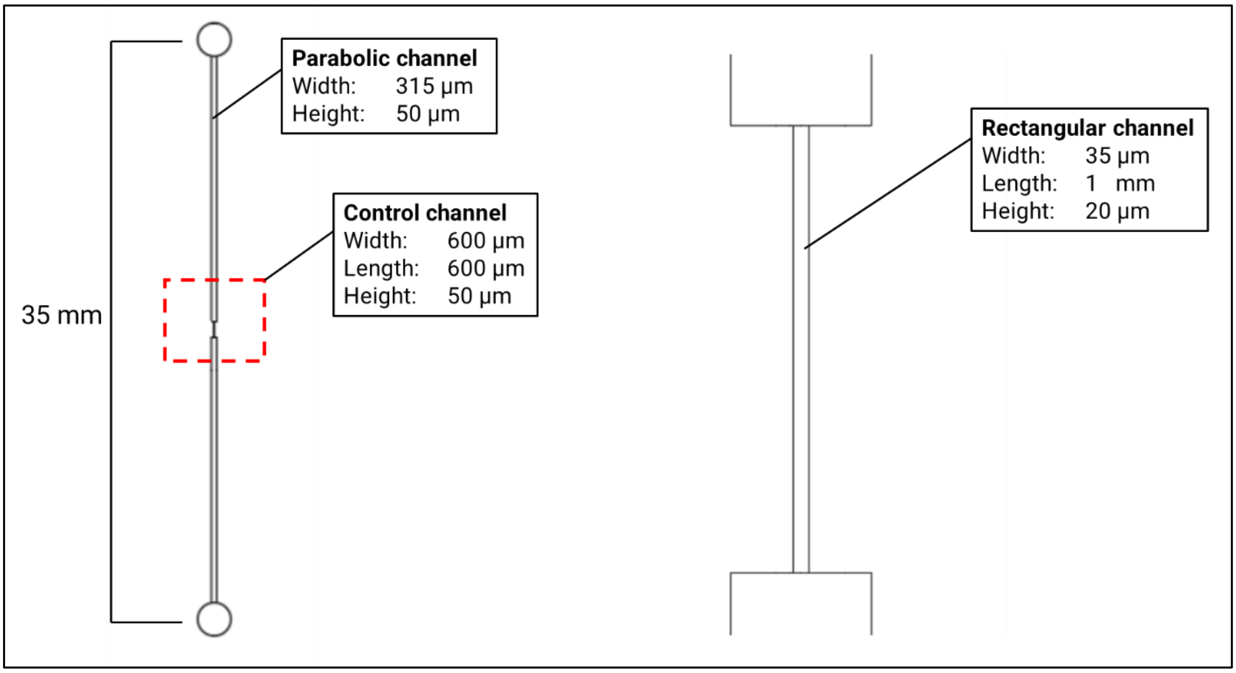


**Figure S1**: The layout of the ACBC microfluidic device featuring all dimensions used.

### Channel profiles resulting from 3D printed moulds

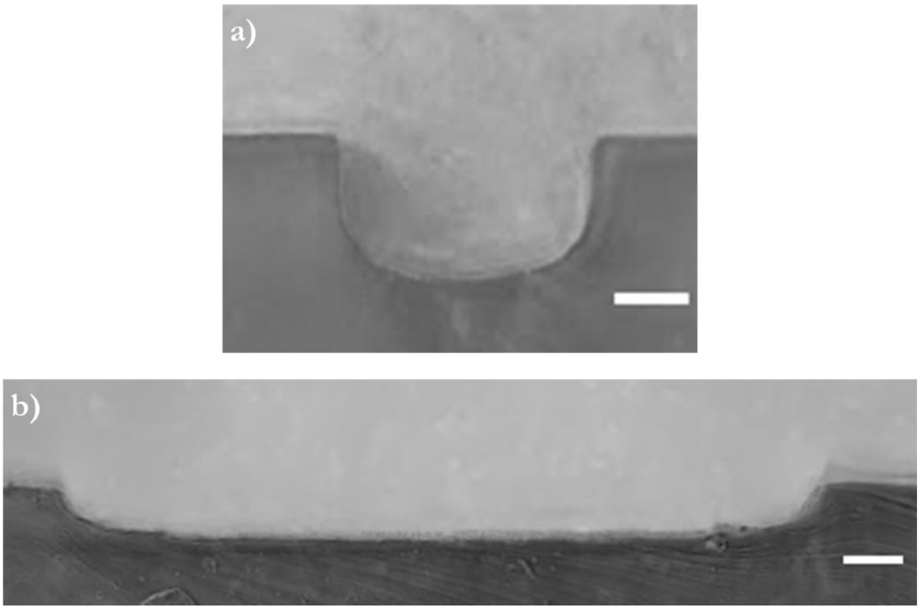


**Figure S2:** Channel cross-section images of the ACBC device. **a)** Cross-section of the rectangular channel (scale bar is 10 µm) and **b)** parabolic channel (scale bar is 30 µm). The angle of the PDMS walls relative to where the substrate is to be placed was determined to be 90º for the narrow channel and 60º for the parabolic channel.

### Fluorescence measurements of adaptive channel height as a function of applied pressure:

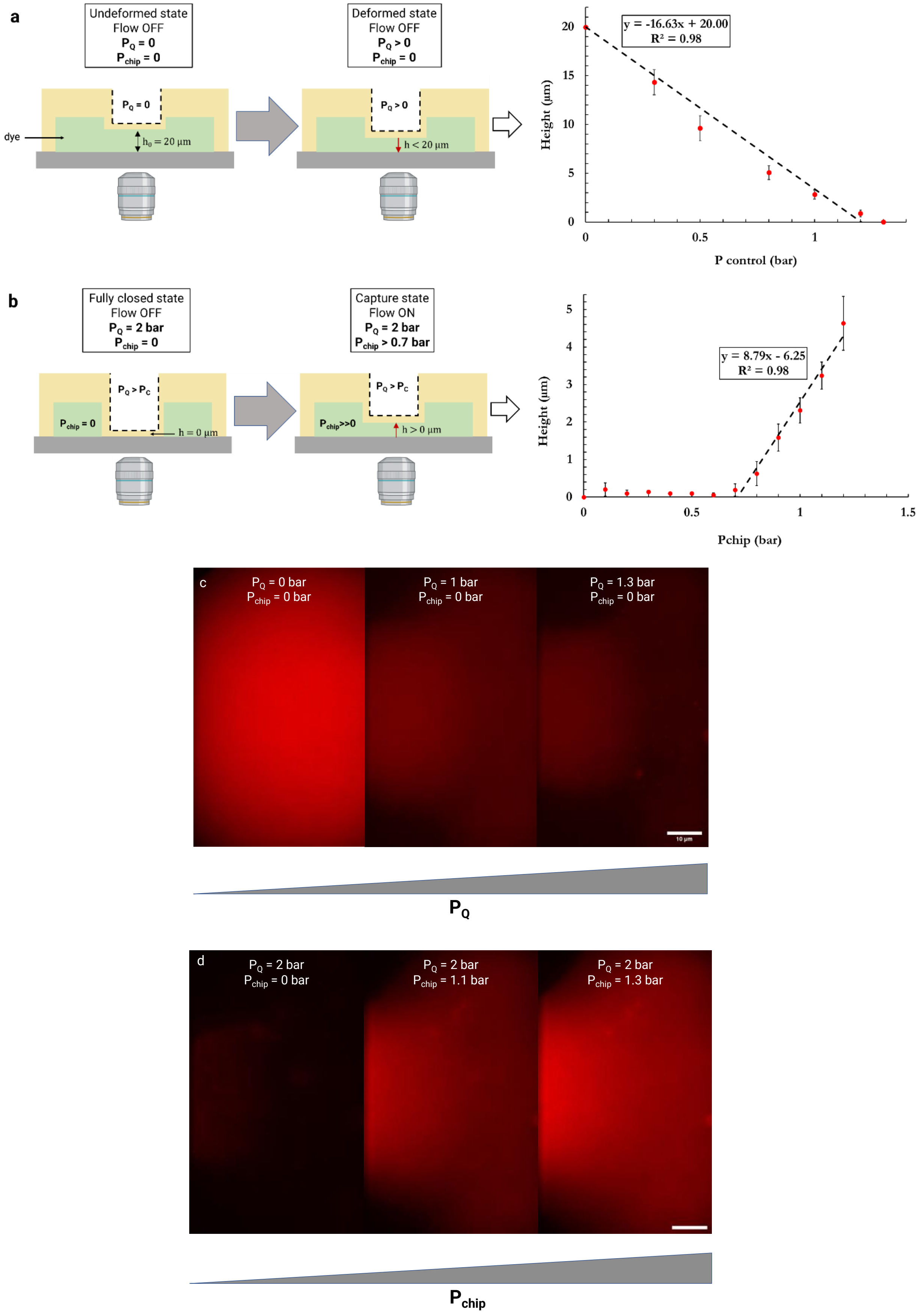

**Figure S3**: Estimation of ACBC channel height using epifluorescence measurements. **a)** 'Close-down' experiments schematic and fluorescence intensity based estimated height of the adaptive channel. The pressure in the control layer is varied with no flow-induced pressure and fluorescence of the dye is acquired. The fluorescence estimates suggest that each bar of pressure changes the channel height by 16.6 µm. **b)** 'Open-up' experiments schematic where the pressure in the control layer is kept constant at 2 bar and flow-induced pressure is varied. It is observed that the height remains collapsed until a pressure of 0.7 bar inside the chip is generated, after which a linear increase in channel height with flow-induced pressure is observed, with a regression coefficient of 8.8 µm/bar. **c)** and **d)** Epifluorescence images for the 'close-down' and 'open-up' experiments at three different conditions. Scale bar is 10 µm.

### Visualization of the capture region

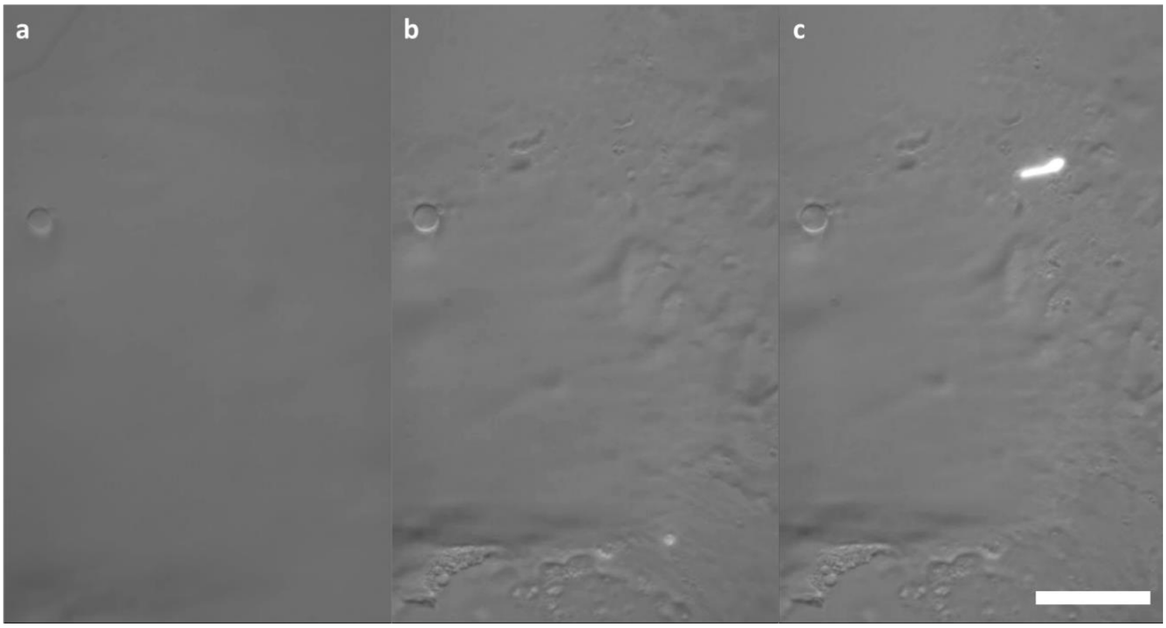


**Figure S4**: Visualization of capture region boundary in HILO mode using 473 nm excitation wavelength. a) The channel is in its original design dimensions. b) The channel is actuated and a capture region is formed. The geometry of the PDMS membrane can be observed. c) An incoming, auto-fluorescent, *E. coli* cell is shown captured in the capture region. The scale bar corresponds to 15 µm.

### Permeabilization assessment: Pathogenic *E. coli* isolate in TEG buffer

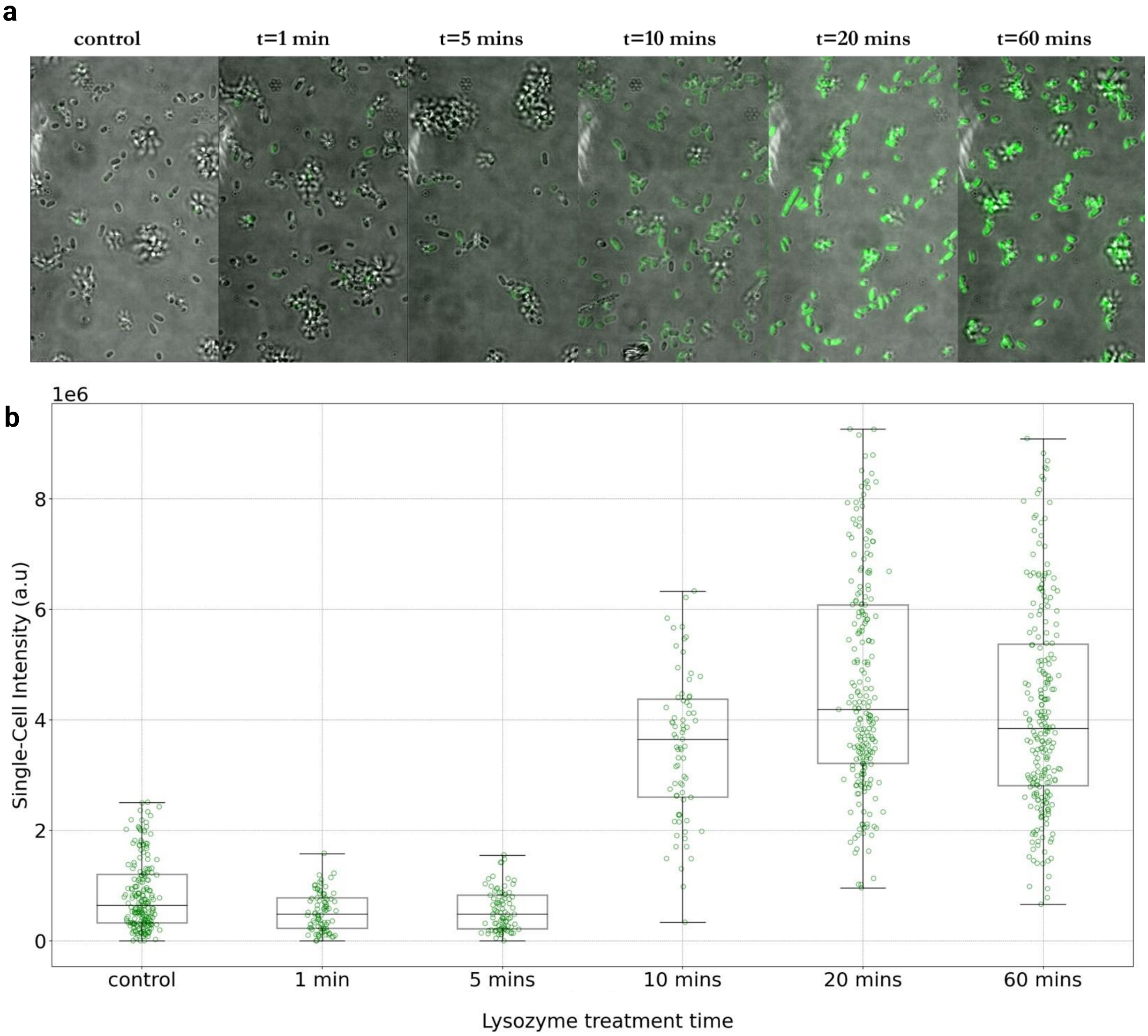


**Figure S5: (a)** Representative images and **(b)** EUB338-Cy3 single-cell intensity boxplots showing the effect of lysozyme incubation time (20 mg/mL, TEG buffer) for a fixed + ethanol treated pathogenic *E. coli* isolate strain. The accessibility of the EUB338-Cy3 probe was used as a proxy to evaluate cell permeabilization as a function of lysozyme treatment time. It appeared that a 20-minute treatment time was sufficient to achieve full permeabilization of this strain. Furthermore, incubation over a longer period (60 minutes) did not appear to have a deleterious effect on 16S rRNA content.

### Fluidic setup used for ACBC chip capture of bacteria.

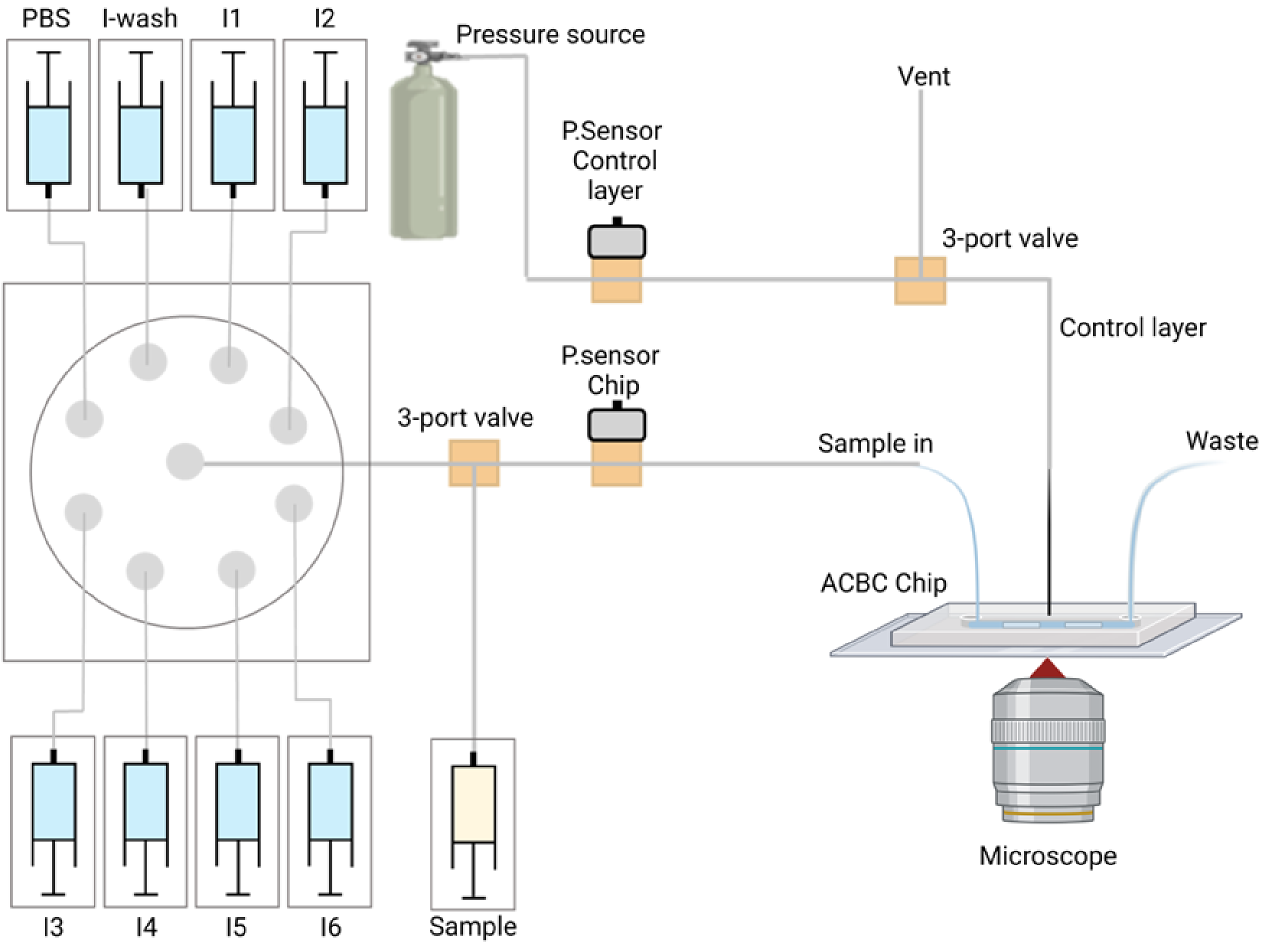


**Figure S6:** Schematic showing the experimental setup used in this study. Reagents are delivered using syringe pumps and a selector valve. Two pressure sensors, one for the control layer and one for the fluidic layer monitor pressure inside channels and enable control of flow-rate as required.

### Trapping of bacteria in a non-adherent ACBC device

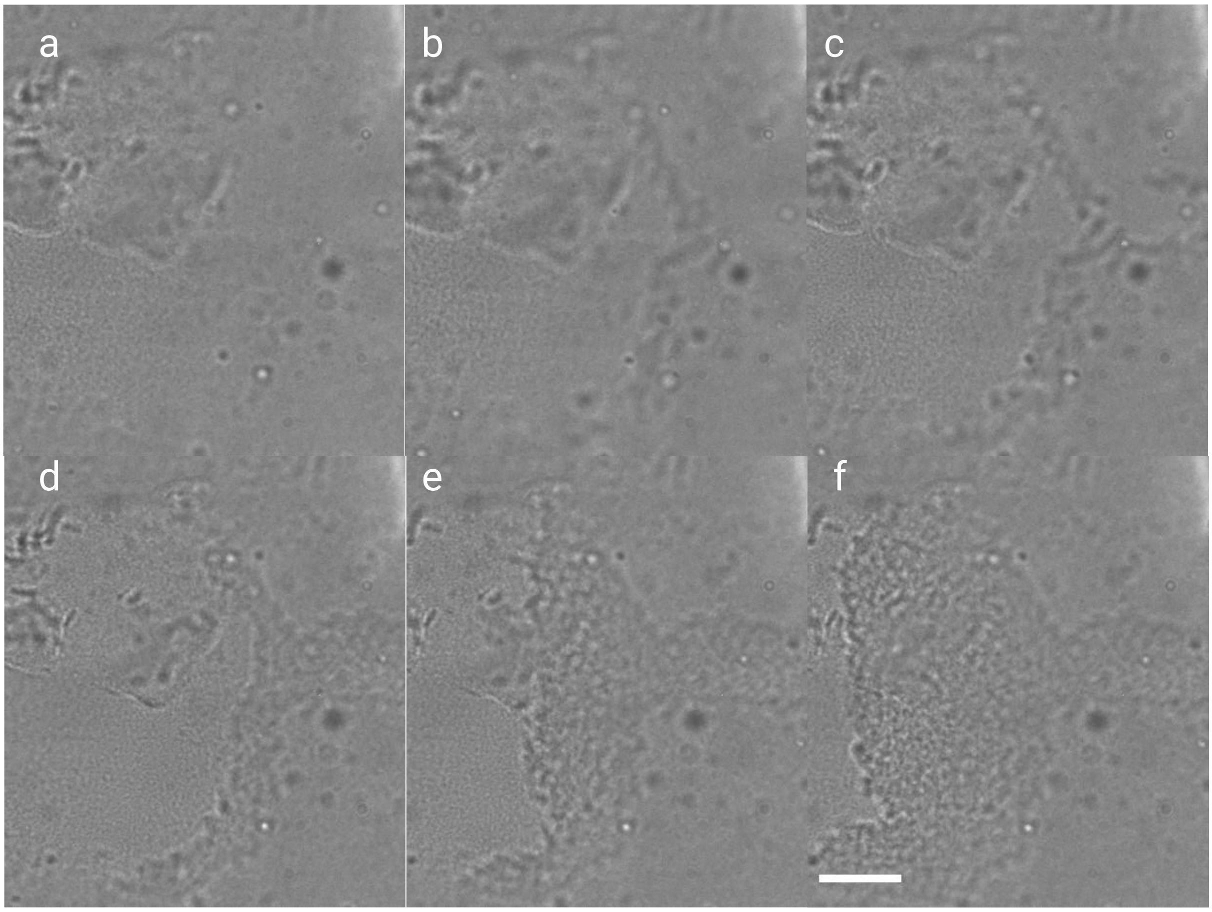


**Figure S7:** Bright-field timelapse of live *E. coli* cell trapping in a PDMS device that was made non-stick towards bacteria in order to visualize the capture region boundary. Cell density for this run was $10^7$ *E. coli* cells/mL over the course of 1 minute. As can be seen all flown cells are trapped at the boundary of the capture region as opposed to being spread evenly at the vicinity of the capture region locus. Scale bar is 10 µm.

**Table S1:** Table containing the encoded FISH probes used for species identification.

| ID | Species | Sequence (5′→3′) | Ref |
|---|---|---|---|
| 1 | *Escherichia coli* | ACCGGTGACGTTAGATTAAAGCTGGGCAAAGGTATTAACTTTACT CCCTTCCTCCCGGGATGCCCCGATTATTGCGTCAT | 1 |
| 2 | *Klebsiella pneumoniae* | ACCGGTGACGTTAGATTAAAGCTGGAGAGCAAGCTCTCTGTGCTA CCGCTCGACTGGTTACATGACTGTCCGTGCTACTC | 2 |
| 3 | *Pseudomonas aeruginosa* | GCCCCTACAGTTCGCAAGTCAGTTAGTTTCCGGACGTTATCCCCCA CTACCAGGCAGTACACCCACCTAGGTCTTGGATG | 1 |
| 4 | *Enterococcus faecalis* | ACCGGTGACGTTAGATTAAAGCTGGCAAGTGTTATCCCCCTCTGAT GGGTAGGTTAGTACACCCACCTAGGTCTTGGATG | 1 |
| 5 | *Streptococcus pneumoniae* | GCCCCTACAGTTCGCAAGTCAGTTACTGGTAGTGATGCAAGTGCA CCTTTTAAGCAGTCTGGTGGACGCAACATTTATAC | 2 |
| 6 | *Streptococcus agalactiae* | GCCCCTACAGTTCGCAAGTCAGTTATCTAGTGTAAACACCAAACCT CAGCGTTCTACGGTTACATGACTGTCCGTGCTACTC | 2 |
| 7 | *Staphylococcus aureus* | ACCGGTGACGTTAGATTAAAGCTGGCATCAGAGAAGCAAGCTTCT CGTCCGTTCGAGTCTGGTGGACGCAACATTTATAC | 1 |

**Table S2:** Table containing the imager FISH probes used for species identification.

| ID | Probe name | Sequence (5′→3′) |
|---|---|---|
| 1 | I1 | **Cy5-**CCAGCTTTAATCTAACGTCACCGGT |
| 2 | I2 | **Cy5-**TAACTGACTTGCGAACTGTAGGGGC |
| 3 | I3 | **Cy5-**GTATAAATGTTGCGTCCACCAGACT |
| 4 | I4 | **Cy5-**CATCCAAGACCTAGGTGGGTGTACT |
| 5 | I5 | **Cy5-**ATGACGCAATAATCGGGGCATCCCG |
| 6 | I6 | **Cy5-**GAGTAGCACGGACAGTCATGTAACC |
| 7 | EUB | **Cy5-**GCTGCCTCCCGTAGGAGT |
